# Three families of CD4-induced antibodies are associated with the capacity of plasma from people living with HIV to mediate ADCC in presence of CD4-mimetics

**DOI:** 10.1101/2024.06.02.24308281

**Authors:** Alexandra Tauzin, Lorie Marchitto, Étienne Bélanger, Mehdi Benlarbi, Guillaume Beaudoin-Bussières, Jérémie Prévost, Derek Yang, Ta-Jung Chiu, Hung-Ching Chen, Catherine Bourassa, Halima Medjahed, Marek K Korzeniowski, Suneetha Gottumukkala, William D. Tolbert, Jonathan Richard, Amos B Smith, Marzena Pazgier, Andrés Finzi

**Affiliations:** Centre de Recherche du CHUM, Montreal, QC, Canada; Département de Microbiologie, Infectiologie et Immunologie, Université de Montréal, Montreal, QC, Canada; Department of Chemistry, School of Arts and Sciences, University of Pennsylvania, Philadelphia, PA, USA; Infectious Diseases Division, Department of Medicine, Uniformed Services University of the Health Sciences, Bethesda, MD, USA

## Abstract

CD4-mimetics (CD4mcs) are small molecule compounds that mimic the interaction of the CD4 receptor with HIV-1 envelope glycoproteins (Env). Env from primary viruses normally samples a “closed” conformation which occludes epitopes recognized by CD4-induced (CD4i) non-neutralizing antibodies (nnAbs). CD4mcs induce conformational changes on Env resulting in the exposure of these otherwise inaccessible epitopes. Here we evaluated the capacity of plasma from a cohort of 50 people living with HIV to recognize HIV-1-infected cells and eliminate them by antibody-dependent cellular cytotoxicity (ADCC) in the presence of a potent indoline CD4mc. We observed a marked heterogeneity among plasma samples. By measuring the levels of different families of CD4i Abs, we found that the levels of anti-cluster A, anti-coreceptor binding site and anti-gp41 cluster I antibodies are responsible for plasma-mediated ADCC in presence of CD4mc.

**IMPORTANCE:** There are several reasons that make it difficult to target the HIV reservoir. One of them, is the capacity of infected cells to prevent the recognition of HIV-1 envelope glycoproteins (Env) by commonly-elicited antibodies in people living with HIV. Small CD4-mimetic compounds expose otherwise occluded Env epitopes, thus enabling their recognition by non-neutralizing antibodies. A better understanding of the contribution of these antibodies to eliminate infected cells in presence of CD4mc could lead to the development of therapeutic cure strategies.

## INTRODUCTION

The human immunodeficiency virus type 1 (HIV-1) envelope (Env) protein is a trimer of gp120/gp41 heterodimers non-covalently associated and highly glycosylated (1, 2). Env is the sole viral protein expressed at the surface of virions and infected cells and mediates viral entry into target cells. During the entry process, Env undergoes different conformational changes transitioning from its native unliganded “closed” conformation (State-1) (3, 4) to a “partially open” conformation (State-2) upon interaction with a single CD4 receptor to an “open” conformation (State-3) after engagement of its three gp120 subunits to CD4 (5, 6). After interaction with the CCR5 or CXCR4 coreceptor (7, 8) Env releases its gp41 fusion peptide which inserts into the cell membrane, thereby triggering fusion between the virion and the target cell (7).

After infection, the HIV-1 accessory proteins Nef and Vpu prevent CD4 expression at the surface of infected cells via different mechanisms. Nef acts on CD4 expressed at the surface of infected cells while Vpu acts on newly-synthetized CD4 in the endoplasmic reticulum (ER), leading to lysosomal degradation or ER-associated protein degradation (ERAD), respectively (9, 10). Thus, Env expressed at the surface of infected cells are free of CD4 and therefore samples a “closed” conformation, preventing recognition of the infected cells by commonly elicited CD4-induced (CD4i) non-neutralizing antibodies (nnAbs). These antibodies were shown to eliminate infected cells by antibody-dependent cellular cytotoxicity (ADCC) when Env samples the “open” conformation (11, 12).

Based on these observations, small CD4-mimetic compounds (CD4mcs) have been used to force Env to adopt more “open” conformations, successfully sensitizing HIV-1-infected cells to ADCC mediated by CD4i Abs and plasma from PLWH (13–17). CD4mcs are HIV-1 entry inhibitors that bind the Phe43 cavity, a highly conserved region of the gp120 (18–21). Binding of CD4mcs to the Phe43 cavity induces conformational changes that allow the exposition of vulnerable CD4i Env epitopes (14, 16, 17, 22–25). To our knowledge, the recently-described indoline CD4mc CJF-III- 288 is the most potent CD4mc to sensitize HIV-1-infected cells to ADCC mediated by PLWH plasma (23). We previously reported the role of two families of nnAbs, recognizing the coreceptor binding site (anti-CoRBS Abs) and the cluster A (anti-cluster A Abs) as being important for PLWH plasma to mediate ADCC in presence of CD4mc (14, 17, 24–26). This combination of nnAbs together with the indane BNM-III-170 CD4mc was shown to stabilize a nnAb-vulnerable Env conformation, State-2A (17). Administration of this combination to humanized mice (hu-mice) resulted in a significant decrease in the size of the viral reservoir (25). In addition to these two CD4i Abs, we recently observed that CD4i nnAbs recognizing the cluster I region of the gp41 (anti-gp41 cluster I Abs), can mediate potent ADCC and reduce HIV-1 replication in hu-mice when combined with CD4mc (16).

In this study, we analyzed the capacity of plasma from a cross-sectional cohort of PLWH to recognize HIV-1-infected cells and mediate ADCC in the presence of a CD4mc. We observed a marked heterogeneity which was associated to the levels of anti-cluster A, anti-coreceptor binding site and anti-gp41 cluster I antibodies.

## RESULTS

### CJF-III-288 enhances the capacity of plasma from PLWH to recognize Env at the surface of infected cells and to mediate ADCC activity

We previously demonstrated that CD4mc sensitizes HIV-1-infected cells to ADCC mediated by plasma from PLWH (13, 23). However, whether the time post-infection or ART treatment could impact the capacity of these plasma to mediate ADCC in the presence of CD4mc remain to be determined. We therefore evaluated the capacity of plasma from a cross-sectional cohort of PLWH to recognize and eliminate HIV-1-infected cells by ADCC. This cohort of PLWH was separated into five different groups: four groups were composed of donors who were not under antiretroviral therapy (ART) and collected at different time post-infection: 0-100 days post-infection; 101-180 days post-infection; 181-365 days post-infection and 2-5 years post-infection. The last group was composed of donors under ART collected 2-5 years post-infection. Basic demographic data of the cohort is summarized in Table 1. Briefly, we infected primary CD4+ T cells with the infectious molecular clone (IMC) HIV-1-_JRFL_ and measured the ability of these plasmas to recognize Env expressed at the surface of productively HIV-1-infected cells (CD4^low^ p24+) by flow cytometry. Plasma samples weakly recognized infected cells in the presence of the vehicle (DMSO, Figure 1A). However, upon addition of the indoline CD4mc CJF-III-288, recognition by all plasma was significantly enhanced. This enhanced recognition translated into a marked increase in ADCC activity (Figure 1B), as measured with a previously described FACS-based ADCC assay (11, 13, 27). As expected, recognition of infected cells by plasma samples was significantly associated with their ADCC activity (Figure 1C). Interestingly, we observed a marked heterogeneity in the capacity of tested PLWH plasma to recognize and mediate ADCC in the presence of CD4mc.

**Figure 1.**
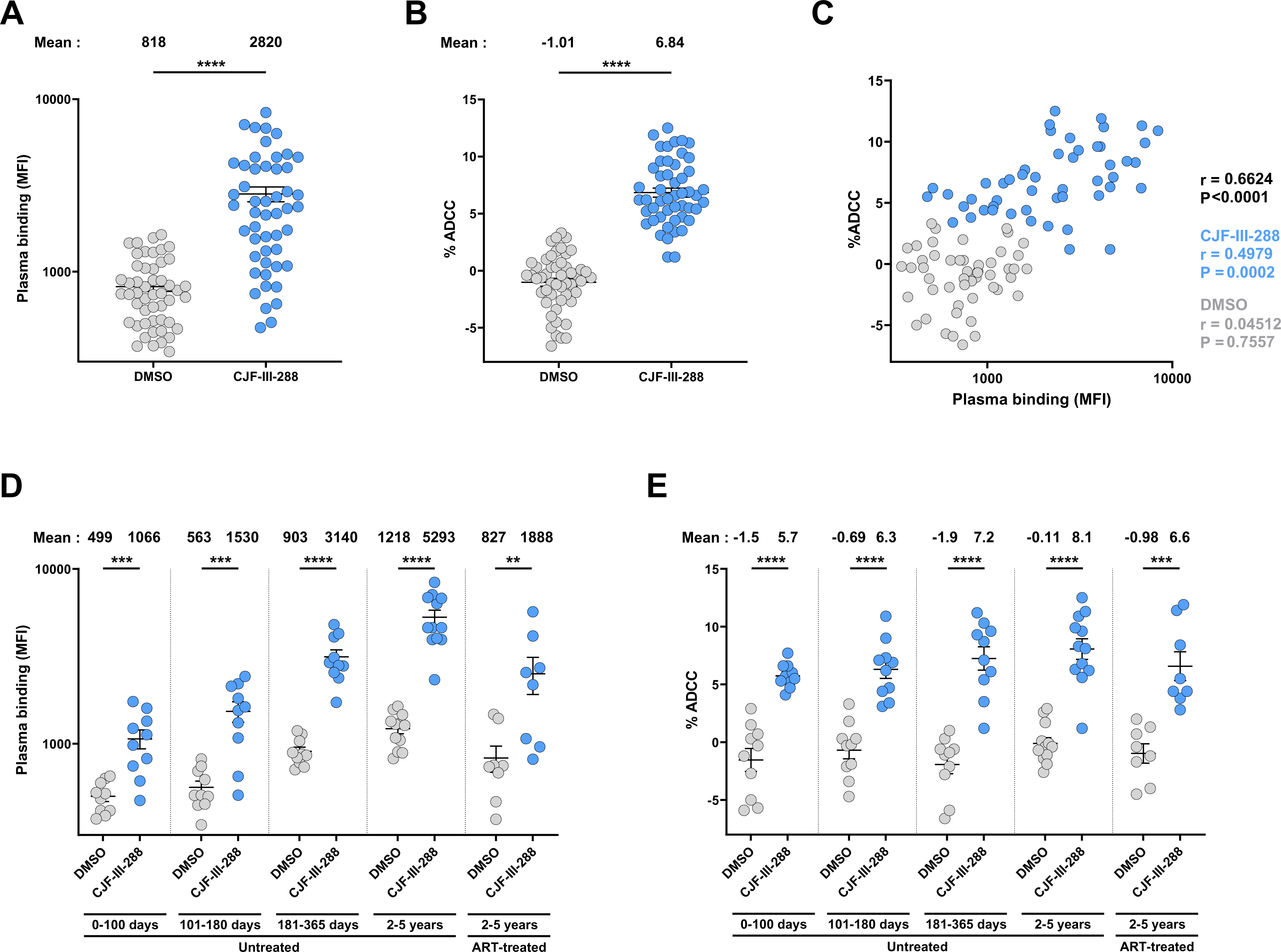
CJF-III-288 enhances recognition and ADCC-mediated elimination of HIV+ infected cells by PLWH plasma. (**A, D**) HIV+ infected CD4+ T cells recognition by 50 PLWH plasma in presence of CJF-III-288 (50 µM) or equivalent volume of DMSO. The panel **A** shows the results obtained with the plasma from the 50 PLWH, and the panel **D** shows the results depending on infection duration and treatment with antiretroviral therapy. (**B, E**) PLWH plasma-mediated ADCC in presence of CJF-III-288 (50 µM) or equivalent volume of DMSO. The panel **B** shows the results obtained with plasma from the 50 PLWH, and the panel **E** shows the results depending on infection duration and treatment with antiretroviral therapy of the individuals. (**C**) Spearman correlation between the capacity of the plasma to recognize the infected cells and to mediate ADCC in presence of DMSO (grey), CJF-III-288 (blue) or both conditions (black). Error bars indicate means ± SEM (∗∗p < 0.01; ∗∗∗p < 0.001; ∗∗∗∗p < 0.0001). The data shows the mean of 3 independent experiments. Statistical significance was tested using paired t tests or Wilcoxon matched-pairs signed rank test, based on statistical normality.

**Table 1:** Characteristics of the cohort of people living with HIV, related to Figures 1, 2 and 3.

| Group | Sex |  | Age<br>(years) | Days since<br>infection | Days between<br>infection and<br>ART initiation | Days since<br>ART initiation | Viral load<br>(copies/mL) | CD4 count<br>(cells/mm<sup>3</sup>) |
| --- | --- | --- | --- | --- | --- | --- | --- | --- |
|  | Female<br>(n) | Male<br>(n) |  |  |  |  |  |  |
| 0-100<br>days | 0 | 10 | 41<br>(18-55) | 68<br>(42-97) | N/A | N/A | 60848<br>(132-391113) | 470<br>(330-829) |
| 101-180<br>days | 0 | 10 | 38<br>(19-58) | 138<br>(109-177) | N/A | N/A | 21663<br>(6641-195302) | 650<br>(420-1235) |
| 181-365<br>days | 0 | 10 | 31<br>(24-44) | 240<br>(203-314) | N/A | N/A | 22785<br>(1866-260852) | 490<br>(230-770) |
| 2-5<br>years | 1 | 11 | 34<br>(20-51) | 1158<br>(856-1810) | N/A | N/A | 292340 (14255-809600) | 410<br>(200-1050) |
| 2-5<br>years | 1 | 7 | 34<br>(21-58) | 1012<br>(792-1612) | 744<br>(19-1375) | 386<br>(192-986) | 50<br>(40-3337) | 585<br>(170-1149) |

\*Values displayed are medians, with ranges in parentheses.

When comparing the responses within plasmas from the different groups of donors, we observed that CJF-III-288 strongly increased Env recognition and ADCC activity compared to DMSO in all groups (Figure 1D-E). We also observed that the capacity of plasma to recognize Env and to mediate ADCC in presence of CJF-III-288 increased with time after infection in untreated donors.

### Three families of CD4i nnAbs contribute to PLWH plasma ability to mediate ADCC in presence of CD4mc

We hypothesized that the differential composition in CD4i nnAbs could potentially explain the observed heterogenicity in the capacity of PLWH plasma to mediate ADCC in the presence of CJF-III-288. Previously, we reported that the combination of anti-CoRBS and anti-cluster A Abs with the indane CD4mc BNM-III-170 can mediate potent ADCC against HIV-1-infected cells both *in vitro* and *in vivo* (14, 17, 24–26). More recently, we reported that anti-gp41 cluster I Abs could also mediate ADCC in the presence of CD4mc (16). To better understand the contribution of these families of CD4i nnAbs to ADCC, we measured their levels respective by indirect ELISA, as described in the material and methods section. All PLWH plasmas had different but detectable anti-cluster A and anti-gp41 cluster I Abs in their plasma (Figure 2A). In contrast, we were unable to detect anti-CoRBS Abs in some donors (Figure 2A). When stratified by groups, only one donor of the 0-100 days post-infection group had detectable anti-CoRBS Abs, and the levels of anti-cluster A Abs were weak compared to the other groups (Figure 2B). In contrast, anti-gp41 cluster I Abs were readily detected in these early samples. The levels of the three families of Abs progressively increased over time and, while the level of anti-gp41 Abs always remained higher than the two other families of Abs, we did not see significant differences between the levels of anti-cluster A and anti-CoRBS Abs after 181 days post-infection. Of note, the capacity of these plasma samples to mediate ADCC correlated with the levels of each of these three families of antibodies in the presence, but not in its absence of CD4mc (Figure 2C-H).

**Figure 2.**
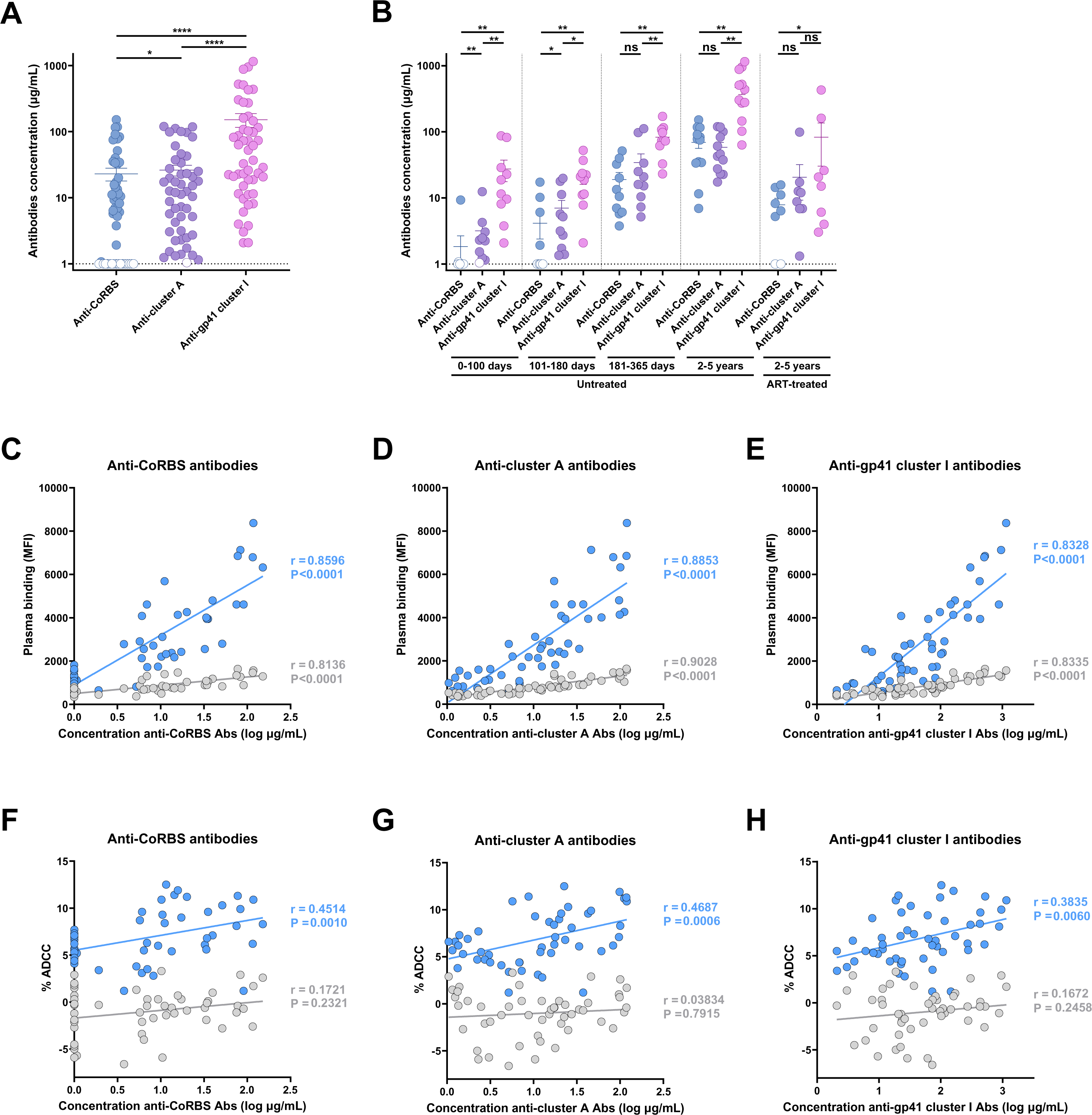
Correlations between the level of three families of non-neutralizing antibodies and plasma binding and ADCC activity. (**A-B**) Indirect ELISA was performed by incubating plasma samples from 50 PLWH to measure the levels of antibodies recognizing specifically the CoRBS, the cluster A or the gp41 cluster I epitopes (see materials and methods section for more details). CD4i Abs was detected using HRP-conjugated anti-human IgG. Relative light unit (RLU) values obtained with BSA (negative control) were subtracted and further normalized to the signal obtained with the appropriate control mAb (17b for the CoRBS, A32 for the cluster A and F240 for the gp41 cluster I ELISA) present in each plate. The panel **A** shows the results obtained with the plasma from the 50 PLWH, and the panel **B** shows the results depending on infection duration and treatment with antiretroviral therapy of the individuals. (**C-E**) Spearman correlations between the capacity of plasma samples from 50 PLWH to recognize the infected cells in presence of DMSO (gray) or CJF-III-288 at 50 µM (blue) and the level of anti-CoRBS (**C**), anti-cluster A (**D**) or anti-gp41 cluster I (**E**) antibodies present in these plasmas. (**F-H**) Spearman or Pearson correlations between the capacity of plasma samples to mediated ADCC in presence of DMSO (gray) or CJF-III-288 at 50 µM (blue) and the level of anti-CoRBS (**F**), anti-cluster A (**G**) or anti-gp41 cluster I (**H**) antibodies present in these plasmas. The data shows the mean of 3 independent experiments. Error bars indicate means ± SEM (∗p < 0.05; ∗∗p < 0.01; ∗∗∗∗p < 0.0001; ns, non-significant). Statistical significance was tested using paired t tests or Wilcoxon matched-pairs signed rank test, based on statistical normality.

To evaluate the contribution of anti-CoRBS, anti-cluster A and anti-gp41 cluster I Abs to plasma-mediated ADCC in the presence of CJF-III-288, we performed a Fab fragment competition assay. Briefly, infected cells were preincubated with CJF-III-288 in presence of anti-CoRBS, anti-cluster A or anti-gp41 cluster I Fab fragments, or a combination of the three Fab fragments. In this experimental setting, Fab fragments bind to their respective epitopes exposed by the CD4mc, and therefore prevent the binding of antibodies from the same family present in plasma from PLWH. This inhibits any ADCC activity mediated by Abs recognizing overlapping epitopes. Preincubation with anti-CoRBS Fab fragments but not anti-cluster A or anti-gp41 cluster I Fab fragments decreased the capacity of PLWH plasma to recognize Env in presence of the CD4mc (Figure 3A). Despite their lack of blocking activity individually, anti-cluster A and anti-gp41 cluster I Fab fragments decreased PLWH plasma binding to infected cells when combined with anti-CoRBS Fab fragments, in agreement with a sequential opening of the trimer by CD4mc in combination with CD4i Abs (14). Interestingly, incubation with each of the three Fab fragments decreased ADCC, albeit this difference was significant only in presence of anti-CoRBS Fab fragments (Figure 3B). Combination of the 3 Fab fragments decreased ADCC levels to those observed in the absence of CD4mc, further demonstrating the importance of these three families of antibodies for the elimination of infected cells by ADCC.

**Figure 3.**
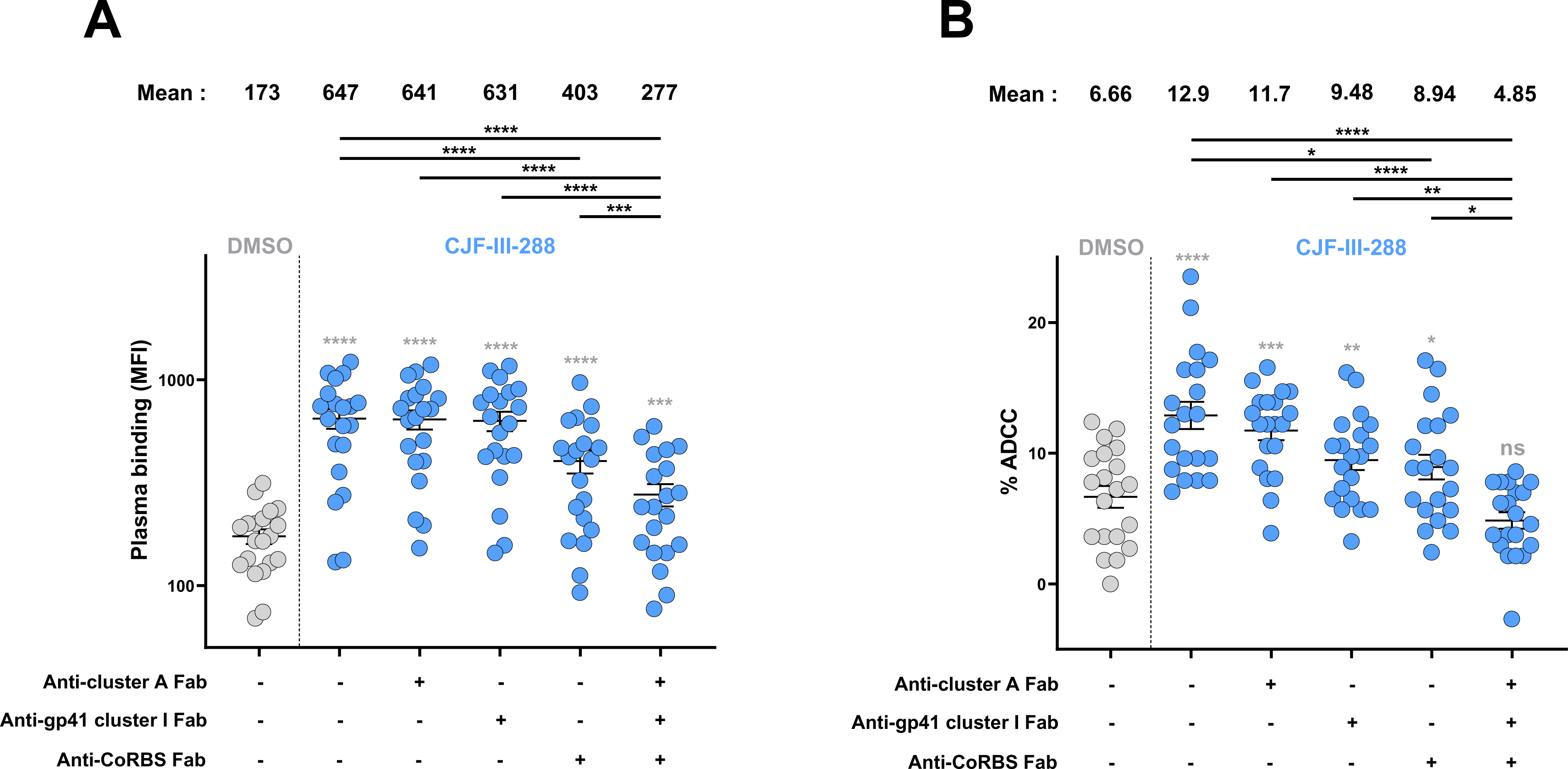
Anti-CoRBS, anti-cluster A and anti-gp41 cluster I CD4-induced Abs contribute to ADCC mediated by plasma from PLWH in presence of a CD4mc. (**A**) HIV+ infected CD4+ T cells recognition and (**B**) ADCC activity of plasma from 20 PLWH (from the two groups of donors, untreated or ART-treated, collected 2-5 years after infection) in presence of DMSO or CJF-III-288 (50 µM, blue) with or without pre-incubation with anti-cluster A A32 Fab, or anti-gp41 cluster I F240 Fab or anti-CoRBS 17b Fab or the three Fab fragments. Error bars indicate means ± SEM (∗p < 0.05; ∗∗p < 0.01; ∗∗∗p < 0.001; ∗∗∗∗p < 0.0001; ns, non-significant). Statistical significance was tested using paired t tests, based on statistical normality.

### Anti-CoRBS, cluster A and gp41-cluster I antibodies contribute to plasma-mediated elimination of autologous primary CD4+ T cells by ADCC

To extend our analysis to primary clinical samples, we expanded CD4+ T cells from different ART-treated PLWH (Table 2) and measured the capacity of each donor’s plasma to eliminate autologous endogenously-infected CD4+ T cells by ADCC in presence of the CJF-III-288 CD4mc. Briefly, CD4+ T cells were isolated from PLWH, activated *ex vivo* with PHA-L/IL-2 and viral replication was followed by intracellular p24 staining (Figure 4A). When we detected at least 7% of p24+ cells in the culture, we evaluated the capacity of autologous plasma to recognize and eliminate infected cells by ADCC in presence or absence of CJF-III-288 (Figure 4B-C). In agreement with our *in vitro* experiments, CD4mc addition significantly increased the recognition of infected cells (Figure 4B), and sensitized them to ADCC (Figure 4C). We then performed Fab fragment competition experiments to evaluate the contribution of anti-CoRBS, anti-cluster A and anti-gp41 cluster I Abs from autologous plasma to mediate ADCC. Preincubation with the combination of the three Fab fragments led to a reduction in ADCC activity, albeit to different extents depending on the donor.

**Figure 4.**
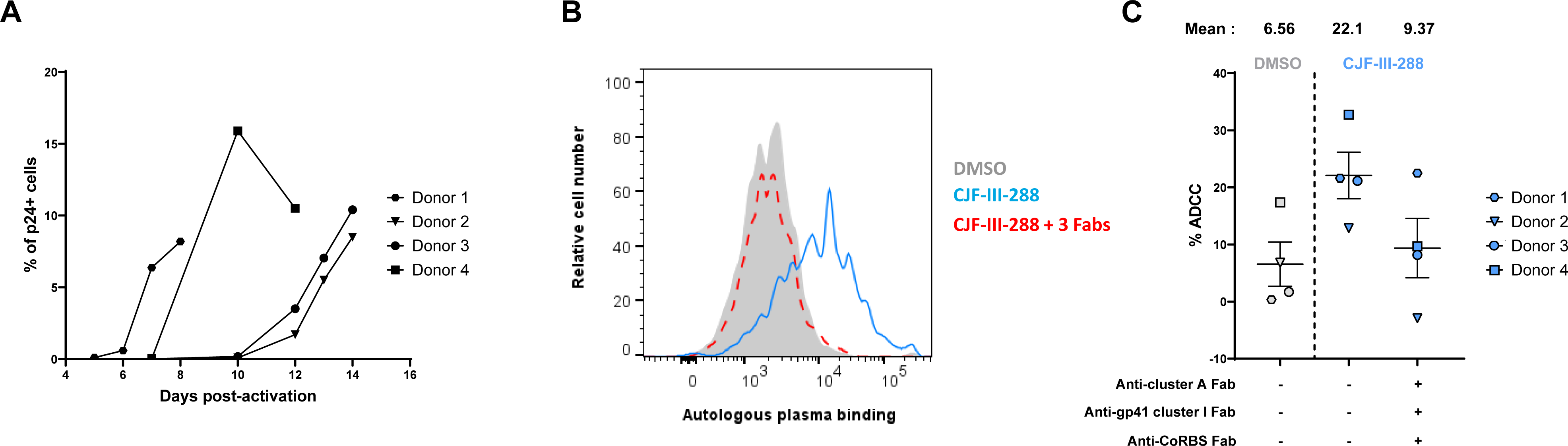
ADCC activity of PLWH plasma against autologous *ex vivo* expanded CD4+ T cells in presence of CJF-III-288. (**A**) HIV+ infected CD4+ T cells isolated from PLWH were expanded *ex vivo*. (**B**) HIV-1-infected cells recognition at the surface of infected cells by autologous plasma in presence of DMSO (grey), CJF-III-288 (blue) or CJF-III-288 with or without preincubation with the three Fab fragments (red). The graph shows the results obtained for a representative donor. (**C**) ADCC activity in presence of DMSO (gray) or CJF-III-288 (50 µM, blue) with autologous plasma after or not pre-incubation with anti-cluster A A32 Fab, anti-gp41 cluster I F240 Fab and anti-CoRBS 17b Fab fragments. Each symbol represent data from an individual donor.

**Table 2:** Demographic characterization of PLWH used to expand primary infected CD4+ T cells, related to Figure 4.

| Donors | Age | Sex | Time since infection (years) | Time since ART initiation (years) | Plasma viral load |
| --- | --- | --- | --- | --- | --- |
| 1-4 | 56 (44-59) | All males | 23 (14-34) | 7.5 (2-13) | All undetectables |

\*Values displayed are medians, with ranges in parentheses.

## DISCUSSION

Env is the only viral antigen expressed at the surface of virions and infected cells, making it the main target of the humoral response (28). However, while antibodies are elicited against Env after HIV-1 infection, only a rare subset of these antibodies - broadly neutralizing antibodies (bNAbs) - effectively recognize Env in its native, “closed” conformation (3, 6). The majority of antibodies commonly induced in PLWH are non-neutralizing and typically recognize regions of Env that are occluded within the unliganded Env trimer (29). Interestingly, epitopes for some nnAbs map to the most conserved regions of Env that are hidden in the unliganded “closed” conformation (18, 30–34). A strategy to harness the inherent potential of these antibodies is to “open-up” Env with CD4mc, thus exposing otherwise hidden epitopes (13, 14, 16, 17). We previously showed that addition of CD4mc sensitize primary infected CD4+ T cells to ADCC mediated by autologous plasma (13). We also showed that CD4mc acts in coordination with anti-CoRBS Abs to “open” Env leading to the exposure of cluster A epitope (14, 24). Anti-cluster A Abs binding stabilizes an asymmetric Env conformation (State-2A) leading to the elimination of infected cells by ADCC (14, 17, 24, 35). Administration of a cocktail of CD4mc with anti-CoRBS/anti-cluster A Abs or with plasma from PLWH results in the significant reduction of the viral reservoir accompanied by a significant delay in viral rebound in hu-mice (25).

In this study, we report that anti-CoRBS Abs, anti-cluster A Abs and a third family of nnAbs (against the gp41 cluster I region), play significant roles in the capacity of any given PLWH plasma to mediate ADCC in the presence of CD4mc. As shown in Figure 5, each of these three families of nnAbs targets highly conserved and non-overlapping regions of Env. Anti-CoRBS Abs interact with regions involved in coreceptor binding (mapping to the outer domain of gp120 at the base of V3 loop), which are necessary for the virus to enter host cells. Since they are involved in coreceptor binding, residues forming the CoRBS are invariable among HIV-1 clades (Figure 5). The cluster A region maps to the gp41 interactive face of gp120, within the topological layers of the gp120 inner domain in the C1-C2 regions, contributing to the structural stabilization of the trimer (33, 36, 37). The cluster A region consists of two non-overlapping sub specificities: A32- and C11-epitope regions (both mapped on Figure 5). The A32 region maps exclusively within the inner domain of gp120 and includes residues within the inner domain mobile layer 1 (centered around CD4 induced α0 helix) of the constant region 1 (C1) region and layer 2 (centered around the α1 helix) of the C2 region. Residues of the gp120 outer domain and the N- and C-termini of gp120 are not part of the A32 epitope. In contrast, the C11 epitope involves residues of the eight-stranded β-sandwich region of the gp120 inner domain that is formed by docking the gp120 N-terminus to the 7-stranded β- sandwich. The gp120 C1, C2 and eight-stranded β-sandwich regions where cluster A epitopes are located are highly conserved across different HIV-1 isolates (Figure 5). During the transition to the CD4-induced conformation, this region undergoes a significant structural rearrangement (18, 30, 32, 33, 38, 39). Finally, anti-gp41 cluster I antibodies recognize a linear epitope in the disulfide loop region (DLR, linking HR1 and HR2 helices) of the principal immunodominant domain (PID) of gp41 (34) (Figure 5). The gp41 sequence recognized by Cluster I Abs 246D and F240 is highly conserved among HIV-1 isolates. F240 has been shown to be broadly cross-reactive and capable of reacting with primary isolates from all clades of HIV-1 (16, 40, 41).

**Figure 5.**
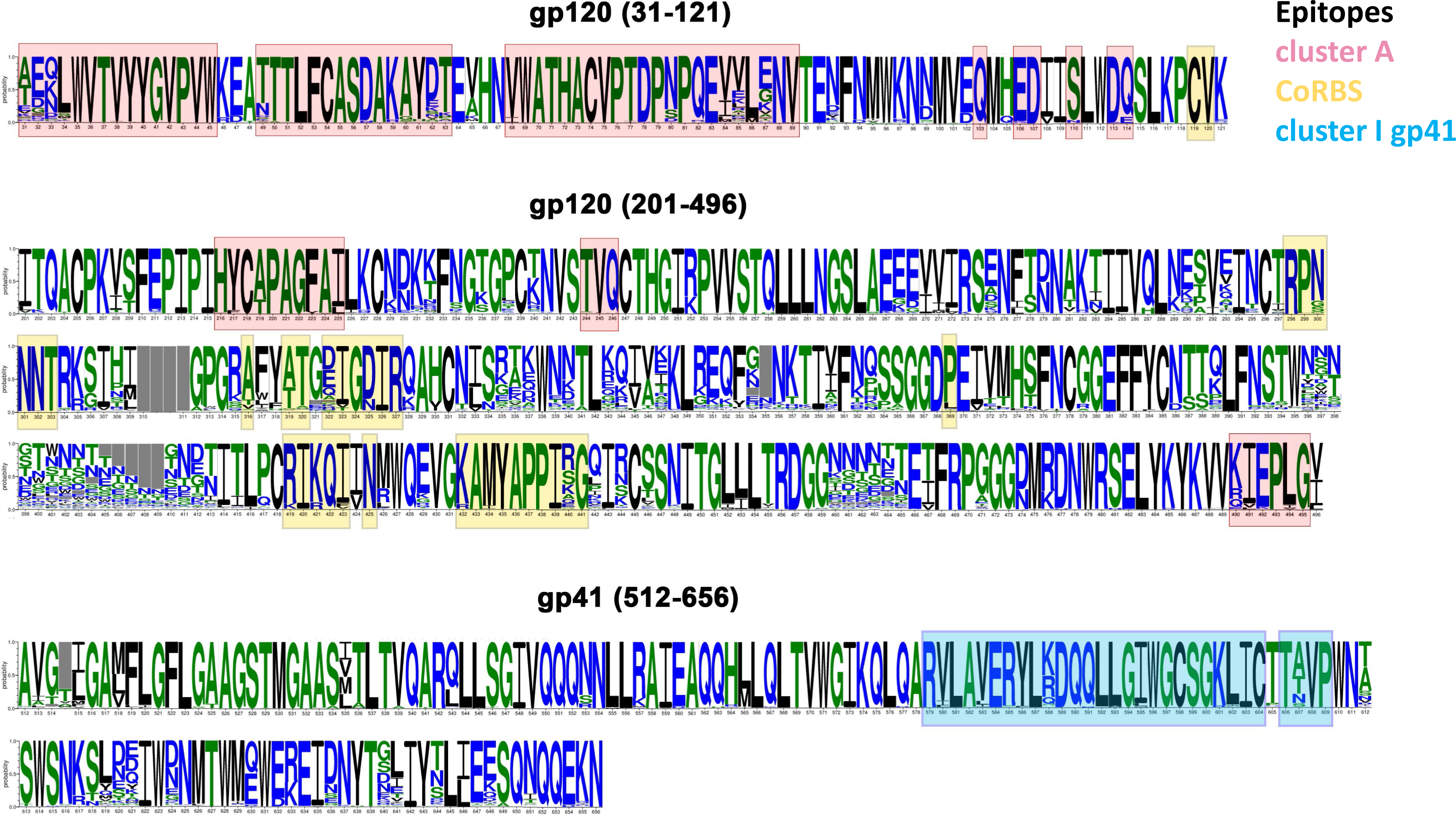
Anti-cluster A, anti-CoRBS and anti-gp41 cluster I Abs recognized highly conserved epitopes in the Env. Env residues forming epitopes for cluster A, CoRBS and gp41 cluster I region are highlighted in pink, yellow and cyan, respectively. Epitope residues are defined as gp120 (cluster A and CoRBS) or gp41 (cluster I) residues contributing buried surface area to the Fab-Env antigen interface as calculated by PISA (http://www.ebi.ac.uk/msd-srv/prot_int/cgi-bin/piserver). Calculations were done based on available Env antigen complexes of cluster A Abs: A32 (PDB code: 4YC2 and 4YBL), N5-i5 (PDB code: 4H8W), 2.2c (PDB code: 4R4F), N60-i3 (PDB code: 4RFO), JR4 (PDB code: 4RFN), CH54 (PDB code: 6MG7), CH55 (PDB code: 6OFI), DH677.3 (PDB code: 6MFP), C11 (PDB code: 6OEJ), and N12-i3 (PDB code: 5W4L); CoRBS Abs: 17b (PDB code: 1GC1), 48d (PDB code: 4DVR), N12-i2 (PDB code: 6W4M), 412d (PDB code: 2QAD), X5 (PDB code: 2B4C); and cluster I Abs: F240 (PDB code: 7N05) and 7B2 (PDB code: 4YDV) Cluster I region includes also 246D epitope residues defined based on peptide mapping as in PMID: 1714520.

Our study emphasizes the importance of these nnAb families in the immune response against HIV- 1, particularly in the presence of CD4mc, which "opens-up" the Env structure, exposing otherwise hidden epitopes and thus facilitating a more effective immune attack on infected cells. This approach holds promise for improving the immune system’s ability to target and eliminate HIV-1- infected cells, potentially contributing to better control of HIV infection. The conserved nature of these epitopes across different HIV-1 strains also suggests that this strategy could be applied cross- clade. Finally, our study suggests that a cocktail comprising monoclonal antibodies against these epitopes, together with a CD4mc, could represent a potent strategy to eliminate HIV-1-infected cells by ADCC.

## MATERIALS AND METHODS

### Ethics statement

Written informed consent was obtained from all study participants and research adhered to the ethical guidelines of CRCHUM and was reviewed and approved by the CRCHUM institutional review board (ethics committee, approval number MP-02-2024-11734). Research adhered to the standards indicated by the Declaration of Helsinki. All participants were adult and provided informed written consent prior to enrollment in accordance with Institutional Review Board approval.

### Cell lines and primary cells

293T human embryonic kidney cells (obtained from ATCC) were maintained in Dulbecco’s Modified Eagle Medium (DMEM) (Wisent, St. Bruno, QC, Canada) supplemented with 5% fetal bovine serum (FBS) (VWR, Radnor, PA, USA) and 100 U/mL penicillin/streptomycin (Wisent). Human peripheral blood mononuclear cells (PBMCs) from five HIV-negative individuals (four males and a female, age range 46-67 years) and four ART-treated HIV-positive individuals (all males, age range 44-59 years) obtained by leukapheresis and Ficoll density gradient isolation were cryopreserved in liquid nitrogen until further use. CD4+ T cells were purified from resting PBMCs by negative selection using immunomagnetic beads per the manufacturer’s instructions (StemCell Technologies, Vancouver, BC) and were activated with phytohemagglutinin-L (PHA-L, 10 μg/mL) for 48h and then maintained in RPMI 1640 (Thermo Fisher Scientific, Waltham, MA, USA) complete medium supplemented with 20% FBS, 100 U/mL penicillin/streptomycin and with recombinant IL-2 (rIL-2, 100 U/mL). All cells were maintained at 37°C under 5% CO2.

### Human plasma samples

The FRQS-AIDS and Infectious Diseases Network supports a representative cohort of newly-HIV- infected subjects with clinical indication of primary infection [the Montreal Primary HIV Infection Cohort (42, 43)]. Plasma samples were isolated from 50 PLWH and separated in five different groups, depending on infection duration and treatment with ART: plasma samples were obtained from never treated individuals during the acute phase of infection (0-100 days after infection), the early phase of infection (101-180 or 180-365 days after infection) or during the chronic phase of infection (2-5 years after infection). Plasma samples were also obtained from a group of chronically infected individuals (2-5 years after infection) under ART treatment. The demographic parameters of the PLWH cohort is described in Table 1.

### Small CD4-mimetic

The small-molecule CD4-mimetic compound CJF-III-288 was synthesized as previously described (23). The compound was dissolved in DMSO at a stock concentration of 10 mM and then diluted to 50 μM in PBS or in RPMI-1640 complete medium for cell-surface staining and for ADCC assays, respectively.

### Antibodies production and purification

FreeStyle 293F cells (Thermo Fisher Scientific) were grown in FreeStyle 293F medium (Thermo Fisher Scientific) to a density of 1 × 10^6^ cells/mL at 37°C with 8% CO_2_ with regular agitation (150 rpm). Cells were transfected with plasmids expressing the light and heavy chains of the anti-cluster A A32, the anti-CoRBS 17b or the anti-gp41 cluster I F240 antibodies using ExpiFectamine 293 transfection reagent, as directed by the manufacturer (Thermo Fisher Scientific). One week later, the cells were pelleted and discarded. The supernatants were filtered (0.22-μm-pore-size filter), and antibodies were purified by protein A affinity columns, as directed by the manufacturer (Cytiva, Marlborough, MA, USA). The recombinant protein preparations were dialyzed against phosphate-buffered saline (PBS) and stored in aliquots at -80°C. To assess purity, recombinant proteins were loaded on SDS-PAGE polyacrylamide gels in the presence or absence of β-mercaptoethanol and stained with Coomassie blue.

The A32, 17b and F240 Fab fragments were prepared from purified IgG (10 mg/mL) by proteolytic digestion with immobilized papain (Pierce, Rockford, IL) and purified using protein A, followed by gel filtration chromatography on a Superdex 200 16/60 column (Cytiva).

### Protein production and purification

ID2 was expressed and isolated as previously described (44). Briefly, stable HEK293 cell lines containing the ID2 expression plasmid were cultured for 6–7 days before the collection of the supernatant and passage through a 0.22 µm filter. Media were then run over an N5-i5 affinity column, washed thoroughly with PBS and protein eluted with 0.1 M glycine at pH3. ID2 was analyzed via SDS-PAGE, dialyzed to PBS and sterile filtered. For gp120 _core_ ΔV1V2V3V5 production, FreeStyle 293F cells (Invitrogen, Carlsbad, Ca, USA) were grown in FreeStyle 293F medium (Invitrogen) to a density of 1 × 10^6^ cells/ml at 37°C with 8% CO_2_ with regular agitation (125 rpm). Cells were transfected with a pCDNA3.1 plasmid encoding codon-optimized His(6)- tagged HIV-1_YU2_ gp120 _core_ ΔV1V2V3V5 using the ExpiFectamine^™^ 293 Transfection Kit as directed by the manufacturer (Invitrogen). One week later, cells were pelleted and discarded. The supernatants were filtered (0.22-μm-pore-size filter) (ThermoFisher Waltham, MA, USA), and the gp120 glycoproteins were purified by nickel affinity columns according to the manufacturer’s instructions (Invitrogen). Fractions containing gp120 were concentrated using Centriprep-30K (EMDMillipore Billerica,MA, USA) centrifugal filter units following the manufacturer instructions. Monomeric gp120 was then purified by FPLC as described (45). Monomeric gp120 preparations were dialyzed against PBS and stored in aliquots at −80°C. To assess purity, recombinant proteins were loaded on SDS-PAGE gels and stained with Coomassie blue.

The gp41 peptide synthesis was previously described (40). The amino acid sequence of the 36- residue peptide of the sequence 583–618 of gp41 (based on the clade B BaL sequence) is gp41_583–618_: VERYLRDQQLLGIWGCSGKLICTTAVPWNASWSNKS. The peptide was synthesized on an ABI 433A automated peptide synthesizer using the optimized HBTU activation/DIEA *in situ* neutralization protocol developed by Kent and colleagues for Boc-chemistry solid phase peptide synthesis (SPPS). After cleavage and deprotection in HF, crude product was precipitated with cold ether and purified to homogeneity by preparative C18 reversed-phase HPLC to afford reduced peptide. The molecular masses were confirmed by electrospray ionization mass spectrometry (ESI-MS). Mass: observed 4082.0 Da, calculated 4081.6 Da. The reduced peptide was dissolved at 0.4 mg/ml in 1 M GuHCl containing 20% DMSO (v/v) for disulfide formation. After 2 hours, reaction was completed and purified with RP-HPLC to afford oxidized peptide. Mass (ESI): observed. 4079.9 Da, calculated. 4079.6 Da.

### Antibodies

The anti-cluster A A32, the anti-CoRBS 17b or the anti-gp41 cluster I F240 antibodies were used as controls in the different ELISA assays. The 17b Fab fragment was used to block anti-CoRBS Abs binding in the anti-CoRBS ELISA assays. Horseradish peroxidase (HRP)-conjugated Ab specific for the Fc region of human IgG (Invitrogen) was used as a secondary Ab to detect Ab binding in ELISA experiments. Alexa Fluor-647-conjugated goat anti-human IgG (Thermo Fisher Scientific) were used as secondary Ab to detect plasma binding in flow cytometry experiments. The A32, 17b and F240 Fab fragments were used to mask the cluster A, CoRBS and cluster I epitopes, respectively, during flow cytometry cell-surface staining and ADCC assays. Anti-human IgG Fc secondary antibodies (Thermo Fisher Scientific) were used when cell surface binding was performed in presence of Fab fragment blockade. The PE-conjugated anti-HIV p24 Ab (Clone KC57-RD1, Beckman Coulter) and the FITC anti-human CD4 OKT4 Antibody (Biolegend) were used to detect productively HIV-1 infected cells in flow cytometry cell-surface staining and ADCC assays (27).

### Plasmids and proviral constructs

The infectious molecular clone (IMC) of HIV-1_JRFL_ was kindly provided by Dr Dennis Burton (The Scripps Research Institute). The vesicular stomatitis virus G (VSV-G)-encoding plasmid was previously described (46).

### Viral production

VSV-G-pseudotyped HIV-1 viruses were produced by co-transfection of 293T cells with the HIV- 1_JRFL_ proviral construct and a VSV-G-encoding vector at a ratio of 3:2 using the polyethylenimine (PEI) method. Two days post-transfection, cell supernatants were harvested, clarified by low-speed centrifugation (300 × g for 5 min), and concentrated by ultracentrifugation at 4°C (100,605 × g for 1h) over a 20% sucrose cushion. Pellets were resuspended in fresh RPMI 1640 complete medium, aliquoted and stored at -80°C until use.

### *In vitro* infections

VSV-G-pseudotyped HIV-1 viruses were used for *in vitro* infection. Briefly, primary CD4+ T cells from HIV-1 negative individuals were isolated from PBMCs, activated for 2 days with PHA-L and then maintained in RPMI 1640 complete medium supplemented with rIL-2. Five to seven days after activation, the cells were spinoculated with the HIV-1_JRFL_ virus at 800 × *g* for 1h in 96-well plates at 25°C. All viral productions were titrated on primary CD4+ T cells to achieve similar levels of infection (around 12% of infected cells).

### *Ex vivo* expansion of HIV-infected CD4+ primary T cells

To expand endogenously infected CD4+ T cells, primary CD4+ T cells obtained from ART-treated PLWH were isolated from PBMCs by negative selection. CD4+ T cells were activated with PHA-L at 10 μg/mL for 48 h and then cultured for several days in RPMI 1640 complete medium supplemented with rIL-2 (100 U/mL). Intracellular anti-p24 staining was performed daily and an ADCC test was done when at least 7% of the cells were p24+.

### Flow cytometry analysis of cell-surface staining

Forty-eight hours after infection, HIV-1-infected primary CD4+ T cells were collected, washed with PBS and transferred in 96-well V-bottom plates. The cells were then incubated for 45 min at 37°C with plasma (1:1000 dilution) in presence of CJF-III-288 (50 μM) or equivalent dilution of DMSO. Cells were then washed twice with PBS and stained with anti-human IgG Alexa Fluor 647- conjugated secondary antibody (2 μg/mL), FITC-conjugated mouse anti-human CD4 (Clone OKT4) Antibody (1:500 dilution) and AquaVivid viability dye (Thermo Fisher Scientific, Cat# L43957) for 20 min at room temperature. Alternatively, for blockade experiments, HIV-1-infected cells were pre-incubated for 15 minutes at room temperature with DMSO/CJF-III-288 and 17b Fab (20 µg/mL), A32 Fab (20 µg/mL) or F240 Fab (40 µg/mL) or the three Fab fragments together before addition of PLWH plasma. Alexa-Fluor-conjugated anti-human IgG Fc secondary antibodies (1:1500 dilution) were used as secondary antibodies. Cells were then washed twice with PBS and fixed in a 2% PBS-formaldehyde solution. The cells were then permeabilized using the Cytofix/Cytoperm Fixation/Permeabilization Kit (BD Biosciences, Mississauga, ON, Canada) and stained intracellularly using PE-conjugated mouse anti-p24 mAb (clone KC57; Beckman Coulter, Brea, CA, USA; 1:100 dilution). Samples were acquired on an Fortessa cytometer (BD Biosciences), and data analysis was performed using FlowJo v10.5.3 (Tree Star, Ashland, OR, USA). The percentage of productively infected cells (p24^+,^ CD4^-^) was determined by gating on the living cell population according to viability dye staining (Aqua Vivid; Thermo Fisher Scientific).

### ADCC assay

ADCC activity was measured using a FACS-based infected cell elimination assay 48 hours after infection. The HIV-1-infected primary CD4+ T cells were stained with AquaVivid viability dye and cell proliferation dye eFluor670 (Thermo Fisher Scientific) and used as target cells. Resting autologous PBMCs, were stained with cell proliferation dye eFluor450 (Thermo Fisher Scientific) and used as effectors cells. The HIV-1-infected primary CD4+ T cells were co-cultured with autologous PBMCs (Effector: Target ratio of 10:1) in 96-well V-bottom plates in the presence of plasma from PLWH (dilution 1:1000) and CJF-III-288 (50 µM) or equivalent dilution of DMSO for 5h at 37°C. For blockade experiments, cells were pre-incubated for 15 minutes at room temperature with DMSO/CJF-III-288 and 17b Fab (20 µg/mL), A32 Fab (20 µg/mL) or F240 Fab (40 µg/mL) or the three Fabs before addition of plasma and effector cells. After the 5h incubation, cells were then washed once with PBS and stained with FITC-conjugated mouse anti-human CD4 (Clone OKT4) antibody for 20 min at room temperature. Cells were then washed twice with PBS and fixed in a 2% PBS-formaldehyde solution. The cells were then permeabilized and stained intracellularly for p24 as described above. Samples were acquired on a Fortessa cytometer (BD Biosciences), and data analysis was performed using FlowJo v10.5.3 (Tree Star, Ashland, OR, USA). The percentage of infected cells (p24^+,^ CD4^-^) was determined by gating on the living cell population according to viability dye staining (Aqua Vivid; Thermo Fisher Scientific). The percentage of ADCC was calculated with the following formula: [(% of p24^+^CD4^-^ cells in Targets plus Effectors) − (% of p24^+^CD4^-^ cells in Targets plus Effectors plus plasma)/(% of p24^+^CD4^-^ cells in Targets) × 100].

### Anti-cluster A ELISA assay

Recombinant stabilized gp120 inner domain (ID2) exposing the A32 epitope was previously described (33, 44). The recombinant proteins were prepared in PBS at a concentration of 0.1 μg/mL and were adsorbed to white 96-well plates (MaxiSorp Nunc) overnight at 4°C. Coated wells were subsequently blocked with blocking buffer (Tris-buffered saline [TBS] containing 0.1% Tween 20 and 2% bovin serum albumin (BSA)) for 1h at room temperature. Wells were then washed four times with washing buffer (TBS containing 0.1% Tween 20). Plasma samples (dilution 1:8000), two-fold serial dilution of anti-cluster A A32 Ab (from 50 to 0.4 ng/mL) to determine concentration using a standard curve, and another dilution of A32 at 1 µg/mL for normalization between plates was prepared in a diluted solution of blocking buffer (0.1% BSA) and incubated with the peptide-coated wells for 90 min at room temperature. Plates were washed four times with washing buffer followed by incubation with HRP-conjugated anti-human IgG secondary Ab (0.3 μg/mL in a diluted solution of blocking buffer [0.4% BSA]) for 1h at room temperature, followed by four washes. HRP enzyme activity was determined after the addition of a 1:1 mix of Western Lightning Plus-ECL oxidizing and luminol reagents (PerkinElmer Life Sciences). Light emission was measured with an LB942 TriStar luminometer (Berthold Technologies). Signal obtained with BSA was subtracted for each plasma and normalized to the signal obtained with A32 present in each plate. Concentration of the plasma was determined with the A32 standard curve. The seropositivity threshold was established using the following formula: mean of 8 plasma from HIV-1-uninfected individuals + (3 standard deviation of the mean of 8 HIV negative plasma).

### Anti-CoRBS ELISA

Recombinant CD4 bound stabilized gp120 proteins, lacking V1, V2, V3 and V5 regions (gp120 _core_ ΔV1V2V3V5 was previously described (25, 33). Proteins were prepared in PBS at a concentration of 0.1 μg/mL and were adsorbed to white 96-well plates (MaxiSorp Nunc) overnight at 4°C. Coated wells were subsequently blocked with blocking buffer (Tris-buffered saline [TBS] containing 0.1% Tween 20 and 2% BSA) for 1h at room temperature. Wells were then washed four times with washing buffer (TBS containing 0.1% Tween 20). Anti-CoRBS 17b Fab fragment (0.5 µg/mL) were prepared in a diluted solution of blocking buffer (0.1% BSA) and incubated with the half of the peptide-coated wells for 90 min at room temperature. The other half of the plate was incubated with a solution of blocking buffer (0.1% BSA). Wells were then washed four times with washing buffer (TBS containing 0.1% Tween 20). Plasma samples (dilution 1:8000), two-fold serial dilution of anti-CoRBS 17b Ab (from 50 to 0.4 ng/mL) to determine concentration using a standard curve, and another dilution of 17b at 1 µg/mL for normalization between plates was prepared in a diluted solution of blocking buffer (0.1% BSA) and incubated with the peptide-coated wells for 90 min at room temperature. Plates were washed four times with washing buffer followed by incubation with HRP-conjugated anti-human IgG secondary Ab (0.3 μg/mL in a diluted solution of blocking buffer [0.4% BSA]) for 1h at room temperature, followed by four washes. HRP enzyme activity was determined after the addition of a 1:1 mix of Western Lightning Plus-ECL oxidizing and luminol reagents (PerkinElmer Life Sciences). Light emission was measured with an LB942 TriStar luminometer (Berthold Technologies). Signal obtained with BSA was subtracted for each plasma and was then normalized to the signal obtained with 17b present in each plate. Concentration of the plasma was determined with the 17b standard curve. The anti-CoRBS antibody level corresponds to the value obtained when the concentration obtained with 17b Fab preincubation condition is subtracted from that without 17b Fab preincubation condition. The seropositivity threshold was established using the following formula: mean of 8 plasmas from HIV-1-uninfected individuals + (3 standard deviation of the mean of 8 HIV negative plasma).

### Anti-gp41 cluster I peptide ELISA

Peptides corresponding to the gp41 C-C loop region (residues 583–618) were either previously described (16, 40) or purchased from Genscript (Piscataway, NJ, USA). Peptides were prepared in PBS at a concentration of 0.1 μg/mL and were adsorbed to white 96-well plates (MaxiSorp Nunc) overnight at 4°C. Coated wells were subsequently blocked with blocking buffer (Tris-buffered saline [TBS] containing 0.1% Tween 20 and 2% BSA) for 1 h at room temperature. Wells were then washed four times with washing buffer (TBS containing 0.1% Tween 20). Plasma samples (dilution 1:8000), two-fold serial dilution of anti-gp41 cluster I F240 Ab (from 50 to 0.4 ng/mL) to determine concentration using a standard curve, and another dilution of F240 at 1 µg/mL for normalization between plates was prepared in a diluted solution of blocking buffer (0.1% BSA) and incubated with the peptide-coated wells for 90 min at room temperature. Plates were washed four times with washing buffer followed by incubation with HRP-conjugated anti-human IgG secondary Ab (0.3 μg/mL in a diluted solution of blocking buffer [0.4% BSA]) for 1h at room temperature, followed by four washes. HRP enzyme activity was determined after the addition of a 1:1 mix of Western Lightning Plus-ECL oxidizing and luminol reagents (PerkinElmer Life Sciences). Light emission was measured with an LB942 TriStar luminometer (Berthold Technologies). Signal obtained with BSA was subtracted for each plasma and was then normalized to the signal obtained with F240 present in each plate. Concentration of the plasma was determined with the F240 standard curve. The seropositivity threshold was established using the following formula: mean of 8 plasmas from HIV-1-uninfected individuals + (3 standard deviation of the mean of 8 HIV negative plasma).

### Sequences analysis

The LOGO plot was created using the Analyze Align tool at the Los Alamos National Laboratory - HIV database which is based on the WebLogo program (https://www.hiv.lanl.gov/content/sequence/ANALYZEALIGN/analyze_align.html) and the HIV- 1 database global curated and filtered 2019 alignment, including 6,223 individual Env protein sequences (47). The relative height of each letter within individual stack represents the frequency of the indicated amino acid at that position. The numbering of Env amino acid sequences is based on the HXB2 reference strain of HIV-1, where 1 is the initial methionine.

## QUANTIFICATION AND STATISTICAL ANALYSIS

Statistics were analyzed using GraphPad Prism version 10.2.0 (GraphPad, San Diego, CA, USA). Every data set was tested for statistical normality and this information was used to apply the appropriate (parametric or nonparametric) statistical test. Statistical details of experiments are indicated in the figure legends. p values < 0.05 were considered significant; significance values are indicated as *p < 0.05, **p < 0.01, ***p < 0.001, ****p < 0.0001.

## Data Availability

All data produced in the present work are contained in the manuscript

## ACKNOWLEDGMENTS

The authors thank the CRCHUM BSL3 and Flow Cytometry Platforms for technical assistance, Mario Legault from the FRQS AIDS and Infectious Diseases network for cohort coordination and clinical samples. This study was supported by grants from the National Institutes of Health to A.F. (R01 AI148379, R01 AI150322, AI176531); A.F. and M.P., (R01 AI129769, AI174908). Support for this work was also provided by the NIAID-funded ERASE HIV consortium (UM1 AI-164562) to A.B.S. and A.F., by a CIHR Team Grant #422148, a project grant #451304 and a Canada Foundation for Innovation grant #41027 to A.F. A.F. was supported by a Canada Research Chair on Retroviral Entry #RCHS0235 950-232424. M.B. and G.B-B. are the recipients of CIHR doctoral fellowship. E.B. is a recipient of a CIHR master fellowships. J.P. was the recipients of a CIHR doctoral fellowship. A.T. was supported by a MITACs Elevation post-doctoral fellowship. The funders had no role in study design, data collection and analysis, decision to publish, or preparation of the manuscript.

## DISCLAIMER

The views expressed in this manuscript are those of the authors and do not reflect the official policy or position of the Uniformed Services University, the U.S. Army, the Department of Defense, the National Institutes of Health, Department of Health and Human Services or the U.S. Government, nor does it mention of trade names, commercial products, or organizations imply endorsement by the U.S. Government.

